# Proteins and transcriptional dysregulation of the brain extracellular matrix in Parkinson’s disease: A systematic review

**DOI:** 10.1101/2023.03.01.23286630

**Authors:** Wote Rike, Shani Stern

## Abstract

The extracellular matrix (ECM) of the brain is a dynamic structure made up of a vast network of bioactive macromolecules that modulate cellular events. Structural, organizational and functional changes in these macromolecules due to genetic variation or environmental stressors are thought to affect the cellular functions, and may result in disease. Most mechanistic studies to date usually focus on the cellular aspects of diseases and pay less attention to the relevance of the processes governing the dynamic nature of the extracellular matrix on disease pathogenesis. Here in this review, we gathered postmortem brain tissue and induced pluripotent stem cell (iPSC)-related studies from PubMed and Google scholar to identify, summarize and describe common macromolecular alterations in the expression of brain ECM components in Parkinson’s disease (PD). According to proteomic studies, proteins such as collagens, fibronectin, annexins and tenascins were recognized to be differentially expressed in Parkinson’s disease. Transcriptomic studies displayed dysregulated pathways including ECM-receptor interaction, focal adhesion, and cell adhesion molecules in Parkinson’s disease. Limited number of relevant studies were accessed from our search indicating that much work still remains to be done to better understand the roles of the ECM in neurodegeneration and Parkinson’s disease. However, we believe that our review will elicit focused primary studies and thus, support the ongoing efforts of the discovery and development of diagnostic biomarkers as well as therapeutic agents for Parkinson’s disease.

## 1. Introduction

The extracellular matrix (ECM) is a 3-dimensional, intracellularly produced, ubiquitous, and complex macromolecular network of proteins and glycans, which is built up around the cellular components of every tissue (1, 2). The ECM demonstrates great tissue specificity due to its varied compositions and topographies that are formed by a dynamic interaction between the numerous cells in each tissue and the altering milieu (3). Brain ECM networks account for 10-20% of brain volume and constitutes collagenous and non-collagenous proteins, glycoproteins, hyaluronan, and proteoglycans (PGs) (4, 5). The network also binds secreted proteins such as growth factors and is known to interact with numerous cell surface receptors, such as integrins thereby providing biochemical cues that regulate the activities of protein complexes and mediate cell-to-cell communication (6, 7). The complex chemical structure enables the ECM to play a crucial role in regulating the most fundamental behaviors and traits of cells including cellular survival, activity, adhesion and growth, and it also aids in the physical organization of neuronal cells into distinct brain areas (8, 9).

Since the brain’s ECM is structured to support homeostatic processes essential to the survival of terminally differentiated neurons, which are thought to be incapable of regeneration, its components are regulated both spatially and temporally throughout brain development (10, 11). When a person approaches adulthood, the brain ECM no longer contains as much collagen and other fibrillar ECM proteins, but it still serves to anchor different structures and guard against abnormal synaptic remodeling (12, 13). Its components are, however, poorly regulated in disease, and as a result, myriad of ECM changes occur during pathogenesis (2, 11, 14). Over the past few decades, the application of high-throughput technologies have fundamentally advanced our understanding of disease mechanisms (15-25). However, understanding the link between neurons and the surrounding ECM still remains a formidable task (11). Many cellular activities are dominated by proteins, and knowing how these processes are controlled at the protein level is crucial for understanding the underlying molecular causes of diseases (26).

The ECM also has an important role in regulating synaptic function and development (27). As the brain develops postnatally, the composition of the ECM undergoes changes to facilitate this function (28, 29). In early stages of development, the ECM is dynamic and permissive to facilitate neuronal plasticity (30, 31). As the brain reaches the end of this critical period, which is marked by extensive neuronal outgrowth and synaptic refinement, the ECM is remodeled and replaced by an adult form enriched by perineural nets (PNN) (12). The PNNs are located between neurons and glia and act as a physical barrier that inhibits further synapse development (29). The composition of the ECM is regulated by neurons through the secretion of ECM proteases such as matrix metalloproteinases (MMPs), and a disintegrin and Metalloproteinase with Thrombospondin motifs (ADAMTS), which play a crucial role in ECM remodeling (32).

Brain ECM components are synthesized and secreted by both neurons and glial cells. However in the CNS, glial cells act as major regulators of the fate of ECM (33). Microglia, the main immune cells in the brain parenchyma, plays an important role in the homeostasis of the brain ECM (34). Microglia perform phagocytic removal of the ECM components during synaptic remodeling (35). It has been suggested that, during synaptic remodeling cell-to-cell contacts occur between microglial processes and dendrites (36), resulting in phagocytic breakdown and remodeling of the ECM by microglia (37). Accordingly, microglia based clearance or modulation of ECM around each synapse serves as the fuel to support synaptic remodeling (38), whereas microglia’s dysfunction results in aberrant ECM clearance or buildup, which contributes to the pathophysiology of the disease (33). In animal models of PD, regions of neuronal degeneration were found to have increased density of microglia (39). Study on mice has also reported brain injury as a promoter of the interaction between microglia and dendrites and subsequent neurotoxicity (40). Activated microglia also causes blood-brain barrier disruption (41), which could lead to fibroblast infiltration and subsequent ECM breakdown in PD (42). Thus, hemostasis of brain ECM is maintained by variety of factors and needs extensive investigation.

Parkinson’s disease (PD) is an extremely heterogeneous neurodegenerative disorder characterized by the cardinal features of the hallmark presence of bradykinesia, rest tremor, and rigidity (43, 44). These motor signs are often preceded by non-motor manifestations such as constipation, autonomic and olfactory dysfunction, sleep disturbances, depression, and anxiety. The motor dysfunction is due, in a large part, to the loss of dopamine (DA)-containing neurons in the substantia nigra pars compacta (SNc) (45-47). At clinical presentation, it has been estimated that more than 60% of SNc DA neurons have already degenerated, and there is also an 80% reduction in dopamine content in the striatum (45, 46, 48). Although the disease appears multifactorial in origin, it could result from a complex interaction between genetics and environment, and commonly affect older people, coming in second only to Alzheimer’s disease in neurodegenerative diseases (49). As a result of the aging population and the world’s increasing industrialization, which is linked to environmental risk factors, the prevalence of Parkinson’s disease is expected to rise steadily to around 13 million by 2040 (50). The late onset of motor symptoms, after the loss of the majority of dopaminergic neurons, and the lack of any reliable biomarkers is the current diagnostic challenge of PD (51). Thus, besides the etiology and treatment-focused studies, discovery and development of specific biomarkers for early diagnosis is an utmost requirement for this disease.

In 1997, Polymeropoulos et al (52) discovered a mutation in the alpha-synuclein gene, SNCA, and showed, for the first time, the link between PD and genetic mutations. Additional evidence also indicated the importance of SNCA gene copy number variations to the pathophysiology of PD (53). It has now been established that PD patients exhibit neuronal loss in the substantia nigra pars compacta (SNc), and there is also deposition of abnormal α-synuclein early in PD, often forming large aggregates termed Lewy bodies (54, 55). There are different genetic risk factors carrying varying degrees of risk for PD in a largely Mendelian fashion. Mutations in some of the genes – *SNCA, LRRK2, VPS35, PARKIN, PINK1*, and *DJ1* – are highly penetrant causes of the disease while heterozygous mutations in the glucocerebrosidase gene (*GBA*) confers a significant risk for PD (56, 57). According to previous research, PARKIN and PINK1 are both involved in a cellular mechanism that preferentially degrades damaged mitochondria in lysosomes by macroautophagy (58, 59). Mitophagy is hampered by the impairment of these genes, which causes an accumulation of damaged mitochondria. Along with numerous antioxidant defenses, PARKIN also indirectly controls the expression of genes necessary for mitochondrial biogenesis (60). These genetic relationships between mitochondrial biogenesis and degeneration suggest that PD may be caused by dysfunctional mitochondrial turnover.

Understanding of PD pathophysiology was substantially aided by proteomic studies of brain tissue (61-63). Samples from PD patients and PD animal models have displayed damage to macromolecules of intracellular components (64-69). Complex disease mechanisms involving neuroinflammation, oxidative stress and ubiquitin proteasomal system (UPS) are commonly linked with PD (70, 71). Disruption to UPS in turn causes the accumulation of clumps of the misfolded α-synuclein protein, an additional impairment of cellular processes, and ultimately neuronal death (72). Proteins are targets for oxidative alterations during oxidative stress, resulting in specific post-translational modifications and impaired protein function. Despite such crucial role of protein homeostasis in PD pathology and many related studies, there has been no single identified therapeutic target that represent the complete picture of PD development.

The lag behind diagnostic biomarkers and effective treatment can be attributed to unbalanced emphasis given to intracellular components with little attention paid to the brain ECM. Brain ECM is vital for neural plasticity and also known to play important role in neurodegeneration (73, 74). Despite its critical role in regulation of cellular function, a few studies have addressed it in PD (75-78). In addition, only a small number of studies that specifically examined the ECM have been performed so far, and these investigations have shown that PD patients have altered ECM components (79, 80). The fact that research is concentrated on certain areas of the brain may also conceal the method by which the ECM and PD association is understood. As demonstrated in postmortem tissue of AD patients (81), PD may also involve numerous distinct locations, including those parts of the brain that have not been known to be impacted by the disease.. Thus, the precise role of the ECM in PD pathology has been masked by these variables, which necessitate comprehensive evaluation of studies that have been conducted on various brain regions and reported alterations of ECM components in PD.

Here, we aimed to examine the differences in ECM expression and composition between Parkinson’s disease patients and matched healthy controls, summarize key findings, and provide recommendations for the direction of future ECM-related research. To do this, we performed a thorough literature search to pinpoint the ECM proteins and genes that are differentially expressed in Parkinson’s disease. This review will support the ongoing effort of exploring the novel roles of ECM components in PD, and could possibly unlock the molecular mechanism behind an aberrant ECM remodeling in PD. It could also contribute to the identification of therapeutic targets for effective management of the disease.

## 2. Methods

### 2.1. The literature search strategy

All articles published in English were searched in PubMed and Google scholar. The information was extracted from proteomic- and transcriptomic studies that reported differentially expressed ECM-related proteins and genes/biological pathways. A comprehensive literature search was done until 10 February 2023 with the search terms: “Proteomic* AND Parkinson’s disease”, “Parkinson’s disease AND Extracellular matrix”, Transcriptomic* AND Parkinson* disease AND Extracellular matrix”, “Gene expression profiling AND Parkinson’s disease”. Only studies that included postmortem/ brain tissue samples and induced pluripotent stem cells (iPSCs)/neurons with human origin were included. The reference list of all identified studies was also scanned for other potentially relevant studies. Following the search, all identified citations were collated and uploaded into a citation management system. The search was re-run before summarizing the data and additional studies retrieved were also screened for inclusion.

### 2.2. Selection criteria

Initially, two independent reviewers screened and retrieved the articles based on the titles and abstracts and then the full texts of the identified articles were evaluated.

#### 2.2.1. Inclusion criteria

- Proteomics studies
- Genome-wide transcriptomic study
- Information on differentially expressed proteins/genes/pathways related to control conditions,
- Employing samples either from human patients or cell lines of human origin.
- Non-review article

#### 2.2.2. Exclusion criteria

- Studies conducted on nonhuman tissue or cell lines
- Interventional studies
- Literature reviews.

### 2.3. Data extraction and management

Two reviewers independently extracted data from the included studies using a well-structured data extraction format with strict adherence to the inclusion criteria. Extracted information includes: author name, year of publication, number of participants, the status of the study participants (case or control), demographic characteristics (e.g. sex, age), PD type (idiopathic or genetic), sample type (brain tissue or iPSCs), brain region, post-mortem interval, proteomic/transcriptomic method, identified proteins/genes/pathways and regulations. The data extracted by the two reviewers were first compared and then merged into one data sheet. The data extraction form and all extracted data are provided as supplementary file. EndNote X7.5 citation manager (Thomson Reuters, New York, USA) was used to store, organize and manage all the references.

Every protein and gene/genomic pathway that has been reported to be altered was manually collected from both the main text and the supplementary materials. The proteins were organized according to their respective human Uniprot ID. Proteins, genes and biological pathways that are commonly reported (reported by, at least, two articles) to be differentially expressed and other relevant evidences were separately presented in the result part below. A summary table with detailed information of the included articles is presented with the supplementary file.

## 3. Results

### 3.1. Literature search results

The total hits from the database and manual search for proteomic studies was 1,243 research articles. Following a title and an abstract screening, 22 articles were found to contain PD-control comparison and proteomic analysis. After a full text review, only 10 articles were identified to contain an ECM protein-related report and differentially expressed ECM proteins and thus, selected for data extraction. The search for transcriptomic studies identified a total of 1,041 studies. These studies were screened based on the title and abstract resulting in 49 articles. Then the full text was reviewed and 24 articles were eligible for inclusion (Fig. 1).

**Figure 1:**
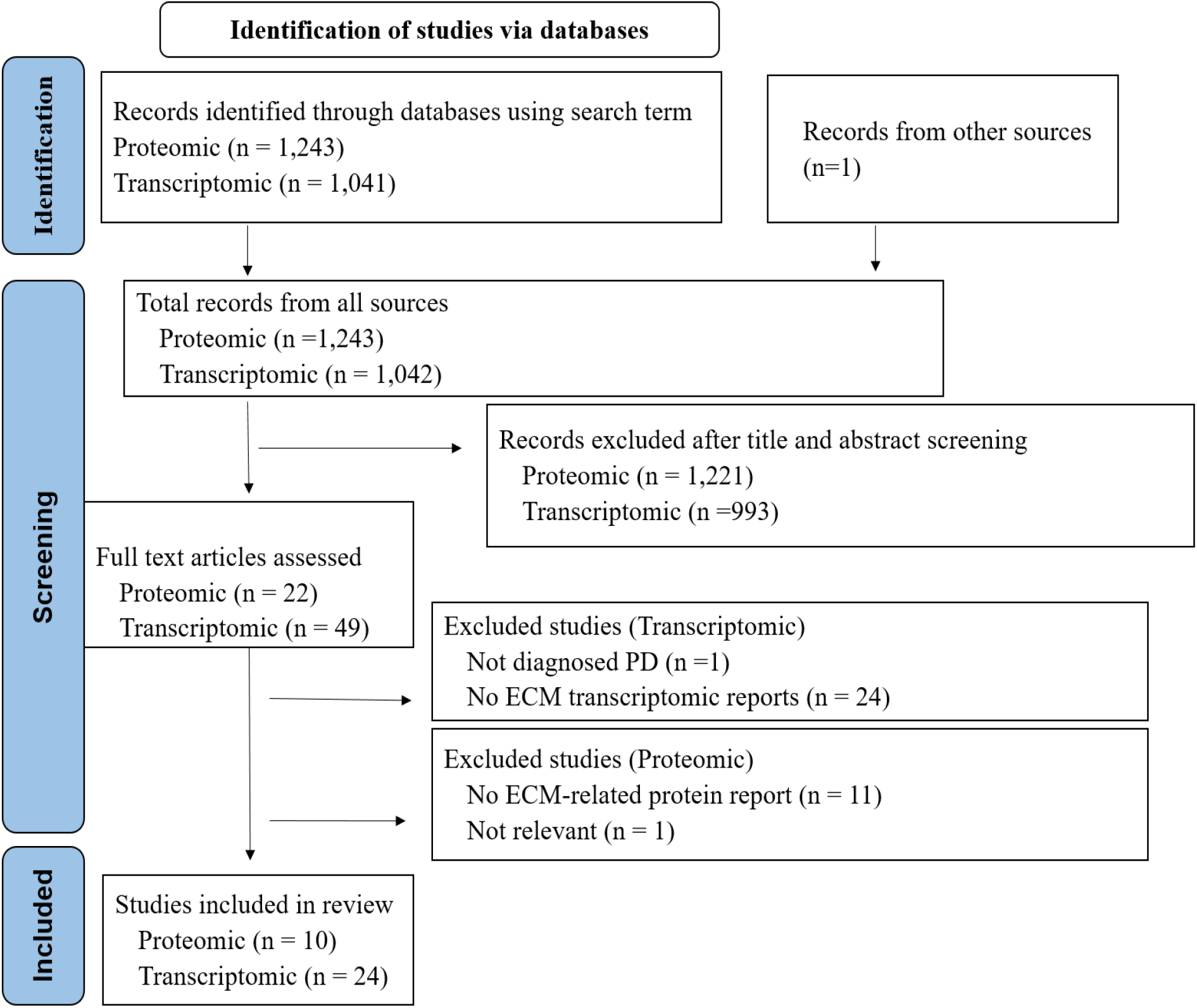
Preferred Reporting Items for Systematic Reviews and Meta-Analyses (PRISMA) flow diagram of the search strategy

### 3.2. Summary of the demographic characteristics of the study participants

All proteomic and the majority of the transcriptomic studies were from post mortem brain tissue-based samples. Four articles with transcriptomic analysis were from iPSCs based studies (15-17, 82). One of the articles reported the analysis from post-mitotic catecholaminergic neuron-like cells, which also constitute DA neurons (83). From the eligible studies, 247 participants (129 cases and 118 controls) were from proteomic analysis and 1,021 participants, (539 cases and 482 controls) were from transcriptomic analysis (Fig. 2). All of the cases in proteomic analysis and most of them in the transcriptomic analysis were idiopathic PD cases (Fig. 3A). The average age of the participants from both proteomic and transcriptomic studies displayed that most of the participants were aged individuals (>65 years) (Fig. 3B). Most studies reported average post-mortem interval (PMI) of less than 22h, and in most of the proteomic studies age, gender and PMI were matched between the case and the controls, if not these variables were controlled (Fig. 3C, Table. 1).

**Figure 2:**
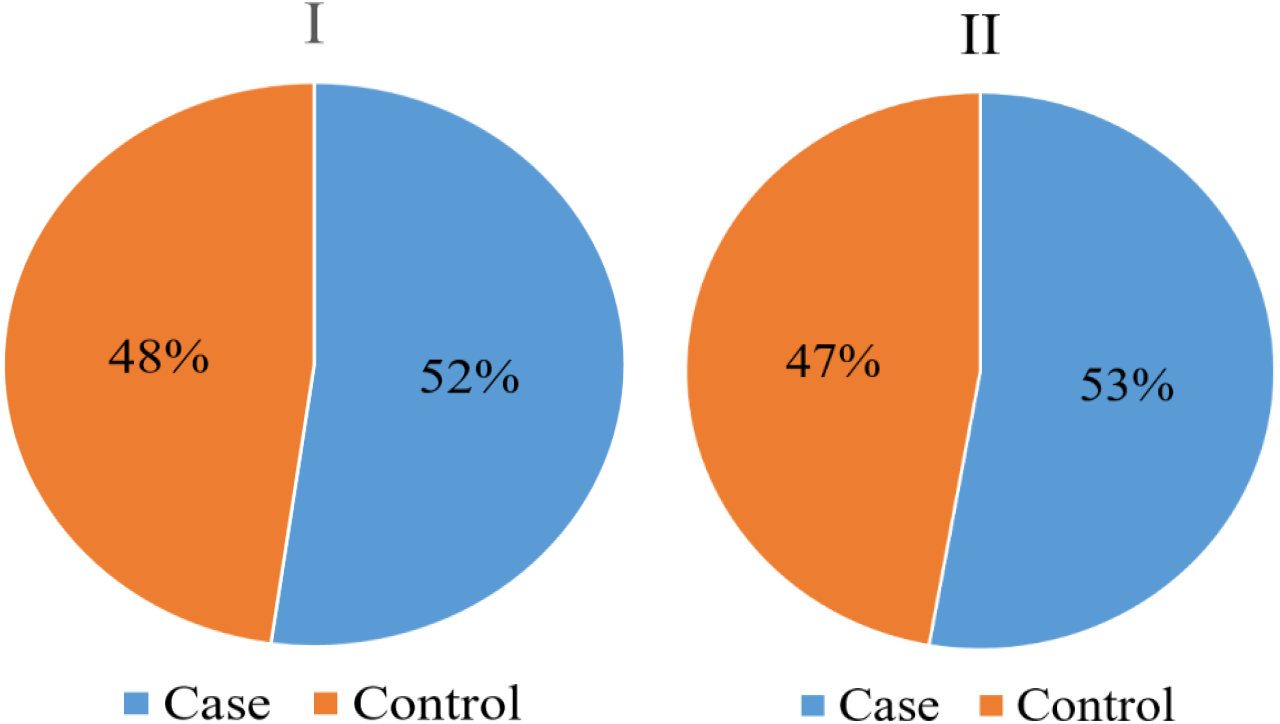
The percentage of PD patients (“Case”) and controls from the included proteomic and transcriptomic studies (I: proteomic, II: transcriptomic)

**Figure 3:**
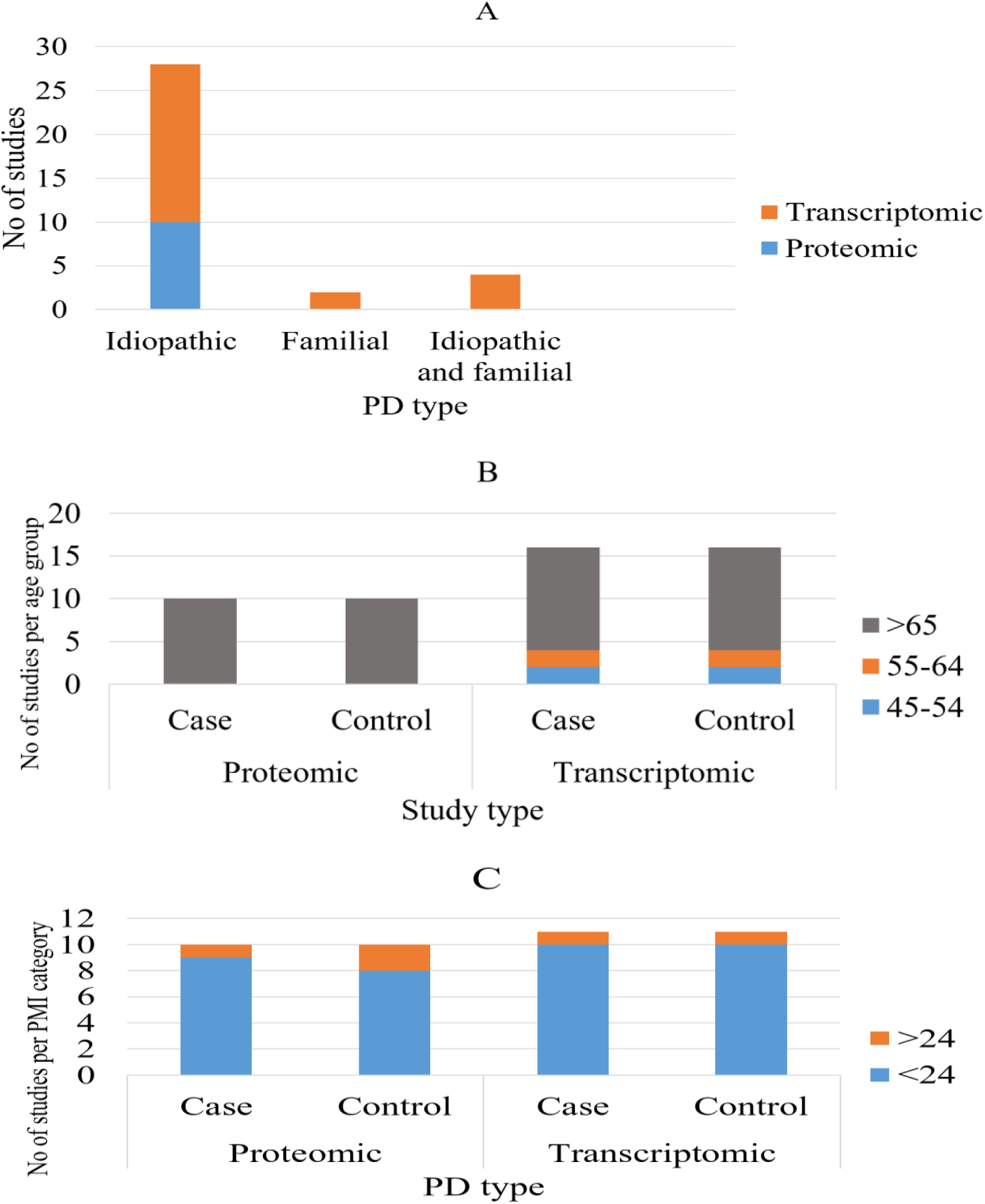
Distribution of patient and sample related variables of both proteomic and transcriptomic studies (A= distribution of PD type; B=Age distribution; C=PMI of brain tissue based studies)

**Table 1:** Patient and sample information from the included proteomic studies

| No of study participants |  | Average age |  | Sex |  | PMI (h) |  | Remark | Ref. |
| --- | --- | --- | --- | --- | --- | --- | --- | --- | --- |
| Case | Control | Case | Control | Male | Female | Case | Control |  |  |
| 6 | 6 | 77.8 | 77.8 | 0 | 12 | 5.75 | 7.15 | Matched* | (84) |
| 3 | 3 | 81.7 | 83.3 | 6 | 0 | 19.7 | 20 |  | (85) |
| 3 | 3 | 79 | 72.7 | 3 | 3 | 18.7 | 24 |  | (86) |
| 20 | 5 | - | - | - | - | <12h | <12h | Matched* | (87) |
| 28 | 37 | 77.6 | 68 | 28 | 37 | 10.25 | 14.1 | Matched*/cohort | (79) |
| 28 | 36 | 77.6 | 68 | 28 | 36 | 10.25 | 14.1 | Matched*/cohort | (80) |
| 5 | 5 | 84.2 | 77.4 | 6 | 4 | 35.6 | 30.2 | Matched*. PMI controlled | (88) |
| 21 | 8 | 79.9 | 77.8 | 17 | 12 | 10.6 | 17.4 |  | (89) |
| 12 | 12 | 76.8 | 79.5 | 24 | 0 | 4.16 | 5.66 | **controlled | (90) |
| 3 | 3 |  |  | 4 | 2 |  |  | Matched* | (91) |

\*Matched = age, gender and PMI are matched; \*\*Controlled = age and PMI are controlled

### 3.3. Description of the included articles

All of the proteomic studies and the majority of the transcriptomic articles were from primary studies of postmortem tissue (Fig. 4 I, Table 2). Except in two studies (79, 86), the reported number of differentially expressed ECM proteins were less than ten. The total share of differentially expressed ECM proteins reported from each proteomic study was less than one fifth of the total differentially expressed proteins in that specific study. The majority of the proteomic studies used samples from the frontal cortex followed by substantia nigra and most of the reported differentially expressed proteins were from these brain regions, too (Fig. 4IV). However, most of the articles in transcriptomic studies incorporated more than one brain region followed by substantia nigra, which was solely used by 37% of the included articles (Fig. 4 II1). Liquid chromatography-tandem mass spectrometry (LC-MS/MS) was the most commonly utilized method for proteomic studies (Tab. 2), whereas microarray with RT-qPCR validation was the common method reported from transcriptomic studies (Fig 4 III)

**Figure 4:**
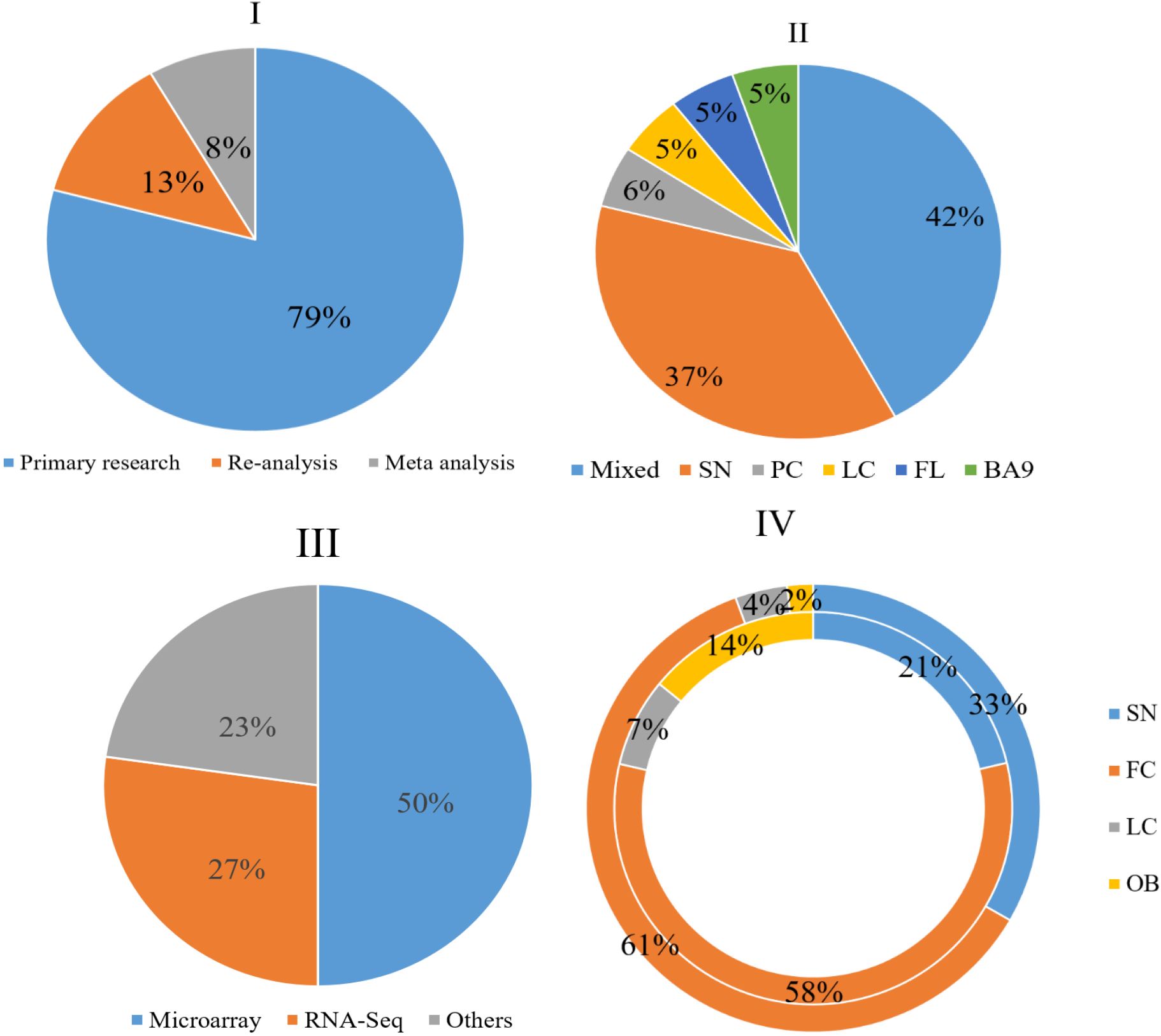
The origin of the included articles (I), Brain region from which the samples were taken for transcriptomic studies (II), methods applied for trancriptomic analysis (III), differentially expressed ECM proteins in comparison with total differentially expressed proteins per brain region (IV). (SN-substantia nigra; FC-frontal cortex; LC-locus ceruleus; OB-olfactory bulbs; FL-frontal lobe; PC -posterior cingulate cortex).

**Table 2:**
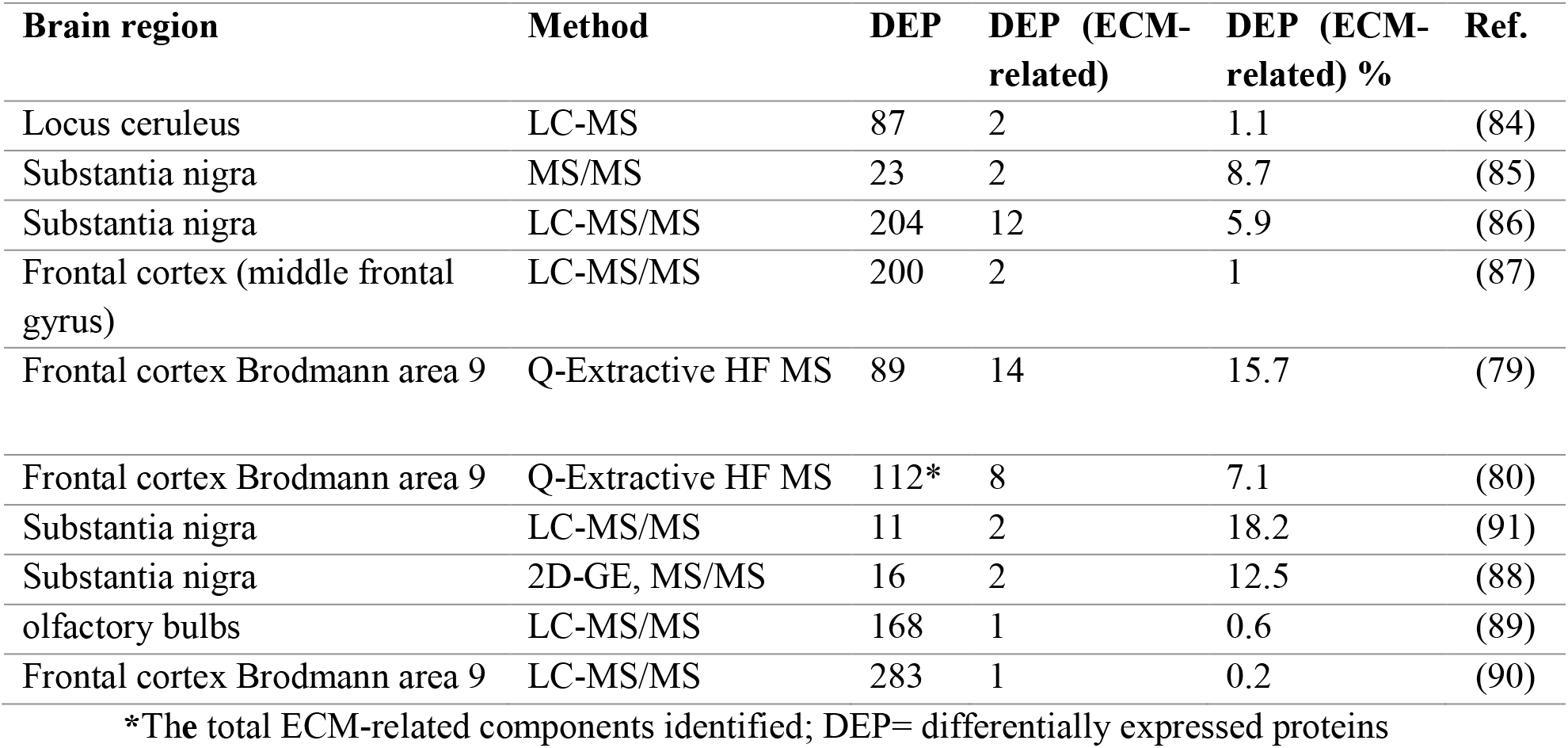
Differentially expressed proteins from postmortem brain samples and the methods applied

**Figure 5:**
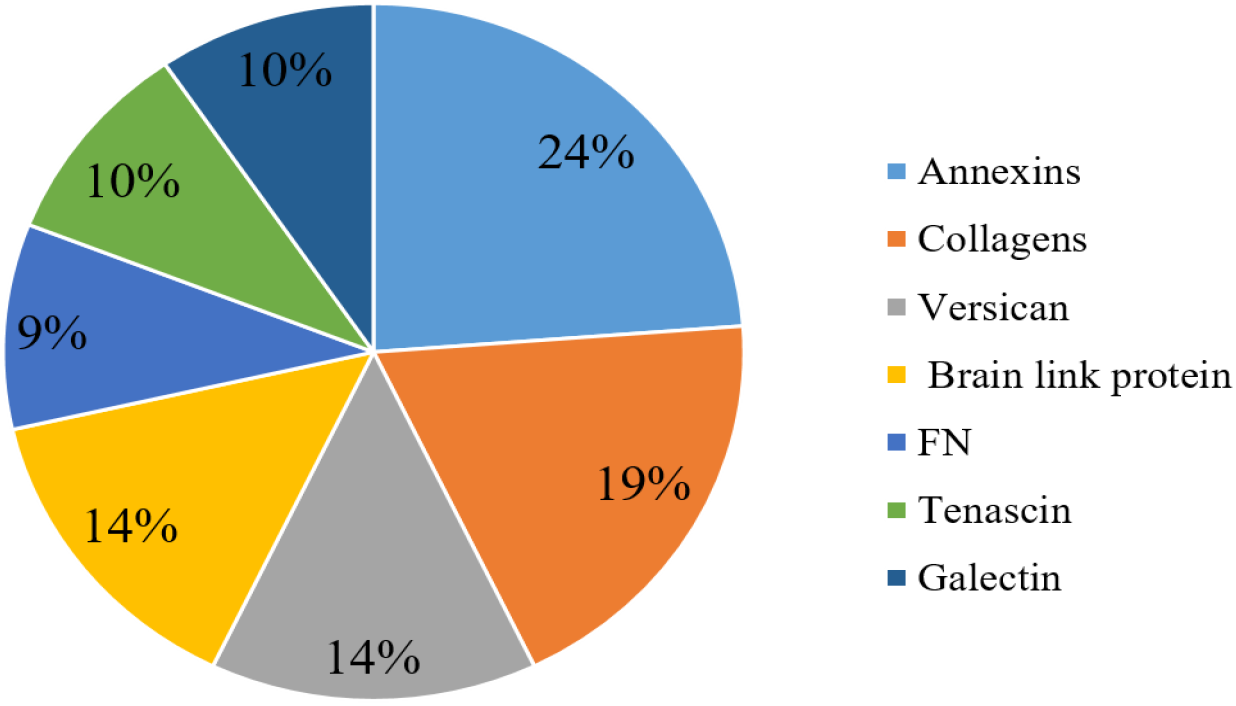
Frequency of commonly reported differentially expressed proteins (FN: Fibronectin)

### 3.4. Description of commonly reported differentially expressed ECM and ECM-related proteins

A total of 46 proteins that are related to the brain ECM were identified from the selected proteomic studies of postmortem tissues. Some of them were reported in more than one study resulting in 33 unique proteins. Annexins and collagens were the most commonly reported proteins followed by versican core protein and brain link protein (Fig.5). Most of the ECM related proteins were reported to be upregulated. All collagen and tenascin subunits were identified to be upregulated. However, different studies reported hyaluronan, proteoglycan link proteins, and fibronectin differently (dysregulated in both directions) (Table 3).

**Table 3:** Commonly reported differentially expressed ECM-related proteins from postmortem tissue

| Components | Protein Names | Gene names | Up/Down | Ref |
| --- | --- | --- | --- | --- |
| Versican family | Versican core protein | VCAN, CSPG2 | up | (79, 80, 84) |
| Collagen family | Collagen alpha-1(I) chain | COL1A1 | up | (79, 80) |
|  | Collagen alpha-2(I) chain | COL1A2 | up | (79, 80) |
|  | Collagen alpha-1(IV) chain | COL4A1 | up | (79) |
|  | Collagen alpha-2(IV) chain | COL4A2 | up | (79, 80, 89) |
|  | Collagen alpha-3(VI) chain | COL6A3 | up | (79, 80) |
| Annexin family | Annexin A1 | ANXA1 | up | (86, 91) |
|  | Annexin A2 | ANXA2 | up | (80, 86) |
|  | Annexin A5 | ANXA5, ANX5 | up | (80, 88) |
|  | Annexin A6 | ANXA6, ANX6 | down | (80, 85) |
| Hyaluronan and proteoglycan link protein family | Hyaluronan and proteoglycan link protein 1 | HAPLN1 (CRTL1) | up | (79) |
|  | Hyaluronan and proteoglycan link protein 2 | HAPLN2 | Down and up | (79, 86, 91) |
|  | Hyaluronan and proteoglycan link protein 4 | HAPLN4 | down | (86) |
| Fibronectin family | Fibronectin (FN) | FN1 (FN) | Up and down | (80, 86) |
| Tenascin family | Tenascin, TN | TNC (HXB) | up | (79) |
|  | Tenascin-R (TN-R) | TNR | up | (87) |
| Galectin family | Galectin-1, Gal-1 | LGALS1 | up | (88) |
|  | Galectin-3, Gal-3 | LGALS3, MAC2 | Down | (84) |
|  | Galectin-3-binding protein | LGALS3BP M2BP | up | (86) |

### 3.5. Description of commonly reported differentially expressed ECM and ECM-related gene groups/ Pathways/ processes

From the transcriptomic studies of postmortem tissues and iPSC-based studies, focal adhesions were the most commonly reported macromolecular assemblies followed by cell adhesion molecules and cell adhesion. ECM-receptor interaction and VEGF signaling were equally reported by three articles (Fig. 6). ECM-receptor interaction, cell adhesion molecules, glycosaminoglycan degradation and integrin signaling were all reported as upregulated across the studies, and collagen and related processes were reported to be downregulated. The remaining gene groups/ Pathways/ biological processes were reported to be dysregulated in both directions. Similar to integrin signaling, all integrin related genes reported in the articles were upregulated, whereas collagen related genes were reported to be dysregulated to both directions (Table 4).

**Figure 6:**
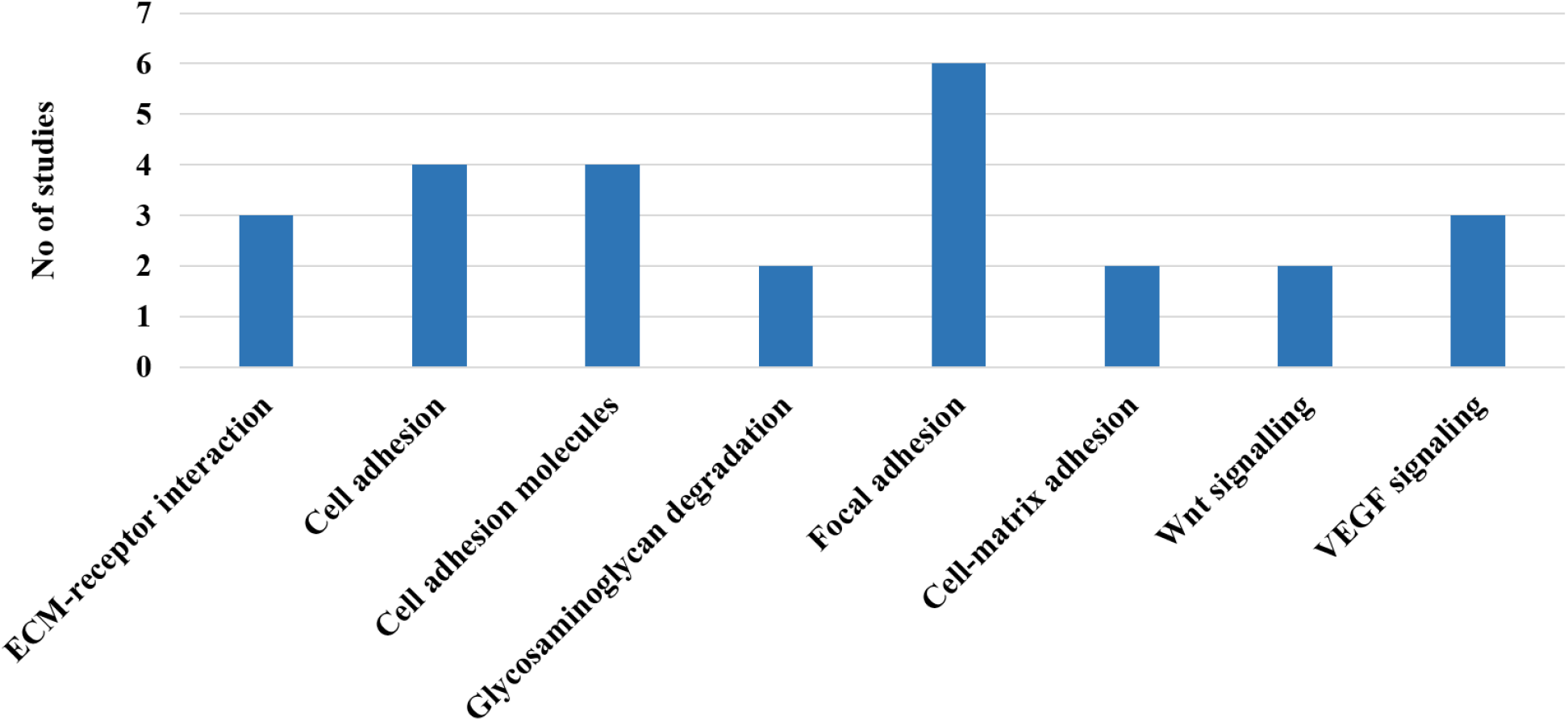
Commonly reported Pathways/ processes/group of genes from postmortem and iPSC-based studies

**Table 4:** Commonly reported differentially expressed ECM-related gene groups/ Pathways/ biological processes

| Genes/ Gene groups/ Pathways/<br>Biological processes | Sample | Up/down | References |
| --- | --- | --- | --- |
| ECM-receptor interaction | Brain tissue, post-mitotic catecholaminergic neuron-like cells, iPSC-derived DA neurons | up | (16, 83, 92) |
| Focal adhesion | Brain tissue and iPSC-derived DA neurons | up | (16, 92-95) |
|  | iPSC-derived DA neurons | down | (15) |
| Cell adhesion molecules | Brain tissue and iPSC-derived DA neurons | up | (15, 92, 94, 96) |
| cell adhesion | Brain tissue | up | (90, 97, 98) |
|  | iPSC-derived DA neurons | Down | (17) |
| Cell-matrix adhesion | Brain tissue | up | (92) |
|  | Brain tissue | down | (99) |
| Glycosaminoglycan degradation | Brain tissue | up | (100, 101) |
| Collagen and related processes | iPSC-derived DA neurons | down | (15, 17) |
| integrin signalling | Brain tissue | up | (90) |
| ITGA1, ITGA 3, ITGA 4, ITGA 5, ITGA 7, ITGA 11, ITGAM, ITGB3BP | Brain tissue and iPSC-derived DA neurons | up | (15, 16, 97) |
| COL1A2, COL4A1, COL4A2 , COL6A3, COL12A1 | iPSC-derived DA neurons | down | (15, 102) |
| COL1A2, COL4A1, COLA2, COL18A1 | Brain tissue and iPSC-derived DA neurons | up | (16, 97) |
| LAMA1, LAMA2, LAMB1, LAMB2 | Brain tissue and iPSC-derived DA neurons | up | (15, 97, 103-105) |
| LAMA3 | Brain tissue | Down | (103) |

## 4. Discussion

The ECM, a collection of cell-secreted molecules, provides biochemical and structural support for the tissues and organs in our body (106). These secreted molecules, particularly the proteins, and cell-bound factors determine structural integrity of the ECM and also allows cells to sense their surroundings via integrin and non-integrin mediated interactions with the ECM (107). These interactions and the interpretation thereof govern the pathophysiologic properties of the cells (106, 108, 109). The alteration of these molecules in the brain in neurodegenerative diseases are reportedly linked (79, 110). Hence, comprehending the molecular dynamics of the ECM offers important insights and helps to uncover ECM signatures linked with PD pathogenesis.

We conducted a comprehensive review of literature to identify ECM proteins and genetic pathways that were differentially expressed in postmortem brain tissue and iPSC-derived neurons. The review included proteomic and transcriptomic studies that reported on ECM changes that occur in PD, which were collected from diverse brain regions. The frontal cortex and substantia nigra were the most frequently sampled regions, with collagens, annexins, tenascins, and versicans being the most commonly reported proteins. The transcriptomic studies identified several differentially expressed ECM-related pathways, including ECM-receptor interaction, focal adhesion, cell adhesion molecules, and cell adhesion. In general, this limited number of studies performed and reporting differentially expressed ECM proteins, coupled with a few ECM-targeted proteomic studies, highlight the need for further work to understand the potential changes of the brain ECM in PD. A lack of adequate ECM targeted studies, as well as an incomplete understanding of its contribution to the pathophysiology of PD, may have hindered the development of effective interventions. The compiled evidence from our review work could potentially enhance our comprehension of the pathophysiology of PD and help the ongoing effort to identify therapeutic targets and develop new treatments.

Around one-third of the ECM is made up of collagens, but there is little data to show how they are affected in PD (111). Despite this barrier, several types of collagens from different categories - fibril-forming (I), network forming (IV) and beaded filament-forming (VI) were observed to be differentially expressed in PD patients compared with matched controls. In our analysis above, we showed a dysregulation of different collagen proteins and genes from brain tissue and iPSC based human studies. Similar findings have also been reported from *in vitro studies*, animal PD models and PD patients. In a 3-D cell culture of primary rat cortical neurons, Cullen et al. (112) observed an association between type IV collagen, the major protein component of the basement membranes, and neurite outgrowth. Transgenic mice with alpha-synuclein overexpression also exhibited elevated type IV collagen expression, implying a potential correlation between alpha-synuclein accumulation and basement membrane dysfunction in PD (113). Type VI collagen is mainly found in the connective compartments of the CNS, and is known to interact with other ECM components (114). An analysis of animal brain sections revealed that the deficiency of collagen VI accelerates neurodegeneration by inhibiting autophagy and inducing apoptosis (115). Additional study on transgenic mice has also demonstrated its neuroprotective role against the toxicity of amyloid-β peptides and UV-induced damage (116). Furthermore, a study on patients with loss-of-function mutations in type VI collagen has also linked this protein to dystonia, a movement disorder characterized by persistent or sporadic muscle spasms (117). Jin et al. (118), in their recent study on sporadic PD patients, reported a possible connection between the COL6A3 gene variants and susceptibility to PD. According to our proteomic review work, collagens IV and VI were found to be upregulated in PD patients (79, 80, 89) although transcriptomic studies showed a dysregulation of Collagen IV in both directions (15, 16). Along with other studies from different PD models, our review work highlights the functional role of collagen (especially Collagen VI) in neuronal cells and their neuroprotective potential against neurodegeneration (119).

The key Perineuronal net (PNN) components, like lecticans, tenascin R, and link proteins, interact with one another to form the PNN’s molecular framework, which wrap around perikaryon and proximal dendrites of certain nerve cells (120). Among lecticans - brevican, neurocan, versican - and other types of proteoglycans like decorin were reported to be differentially expressed in PD in the proteomic studies included in the current review work (79, 80, 84, 86). However, only the versican protein was widely reported among them and observed to be upregulated across the studies (79, 80, 84). Versican is a non-fibrous component of the brain’s ECM, acting as a core protein to which side chains of carbohydrates bind to create proteoglycans (106). It is a multifunctional protein modulating cell adhesion, migration, and inflammation thereby interacting with immune cell receptors and also other ECM components like fibronectin, and tenascin (121-123). The binding of immune cells to the versican-ECM complex may breakdown the ECM leading to neuroinflammation and apoptosis (123, 124). However, whether the intact or fragmented versican is responsible for neuroinflammation and apoptosis needs further investigation. According to Downs et al (80), its alteration involves both proteomic and glycoproteomic changes in PD and they emphasized the importance of changes in its glycosylation pattern on the inflammatory process in PD. Overall, further research into its neuroinflammatory mechanism and targeted work could lead to a novel approach to treating PD.

According to the reports from proteomic studies included in the current review, fibronectin, and tenascin were also among the widely reported dysregulated ECM proteins (79, 80, 86, 87). Tenascins (C and R) were demonstrated to be upregulated but mixed results were observed in the case of fibronectin (80, 86). Such opposing expression of different glycoproteins was also implicated in multiple sclerosis (125). Like in PD, high tenascin level was also reported from the brains of Alzheimer’s disease (AD) patients (120). According to the study conducted on multiple sclerosis patients, it has been suggested that the enhanced production may probably represents a defensive mechanism, yet excessive production could lead to disorganized matrix depletion and the suppression of restorative activities (106, 126). Additional studies on *in vitro* model of induced inflammation of hippocampal neurons co-cultured with glial cells and in an AD mouse model showed that inhibition of its function and compounds that reduce its production suppress neurodegeneration (127, 128). These outcomes in other neurodegenerative diseases and its upregulation in PD patients indicate the importance of tenascins as a potential therapeutic target for neurodegenerative diseases. In order to stabilize the ECM at the cell surface, fibronectin need uninterrupted polymerization into fibrils, which in turn requires adequate delivery of integrins (129, 130). In our literature review the integrin gene expression was upregulated in both postmortem and iPSC-based studies PD patients (15, 16) and opposite expression between fibronectin and integrin was also reported in iPSC based studies (15, 16). Such contradicting results from co-functioning genes necessitate further work to figure out their exact contribution in PD pathogenesis.

Annexins are another group of proteins reported to be differentially expressed in postmortem tissue of PD patients. According to the report from affinity chromatography and solid phase assays, these proteins were known to bind with glycosaminoglycans (GAGs), ECM components, with specific binding affinities (131). In this review work, annexin A6, was downregulated while annexins A1, A2 and A5 were upregulated (80, 86, 88, 91). It has been demonstrated that annexin A6 acts as a recognition component for GAGs in the extracellular space (131). In contrast to annexin A6, transcriptomic postmortem studies from our review work indicated upregulation of the GAG degradation pathway (100, 101). Such opposing expression may likely indicate a disruption of their molecular networks and associated signaling pathways in PD (131). Furthermore, it has been demonstrated that, in conjunction with annexin A2, annexin A6 interacts with tau, which is thought to contribute to the pathological redistribution of tau in Alzheimer’s disease. (132). Recombinant human annexin A1 (hrANXA1) was demonstrated to lower amyloid-β levels in an AD mice model (133) whereas annexin A5, whose cerebrospinal fluid level was reported to match disease severity in AD patients, was implicated as a biomarker in AD (134). These evidences together imply the impact of annexins in neurodegeneration and their potential as biomarkers and therapeutic targets.

In addition to differentially expressed proteins, changes in gene expression levels was reported. A number of pathways and processes, including ECM-receptor interaction, focal adhesion, cell adhesion molecules and cell adhesion were observed to be dysregulated after gene ontology (GO) analysis. Focal adhesions are the specialized cell adhesion structures that mediate the interaction between the ECM and intracellular actin cytoskeleton (135, 136). Cell adhesions occur through the interactions between <u>cell-adhesion molecules</u> (CAMs) and transmembrane proteins located on the cell surface, which connect cells to the ECM (137). These interactions involve two types of receptors, cadherin and integrin receptors, which mediate cell-cell and cell-ECM adhesion, respectively (138, 139). In the ECM, integrin binds with laminins, cell adhesion molecules, and major components of basement membrane (140). From this review work, both integrins and laminins were observed to be upregulated in PD (15, 16, 90, 97, 103-105). Furthermore, the significance of cell adhesion for cell survival and physiology highlights the importance of proper communication between ECM and integrins (141). These evidences, together with dysregulation of their signaling pathways and individual genes in PD, underline integrins as a potential and valid target molecules for PD treatment. Previous success in developing integrin-targeted antibodies thereby blocking ligand binding (142, 143) and downstream signaling (142, 144) would further support the significance of integrin as important drug targets..

According to some of the included studies, the substantia nigra, the primary region of the brain involved in PD, has relatively larger percentage of differentially expressed ECM proteins per total number of differentially expressed proteins compared to other brain regions. Overall, most of the proteomic studies from our review reported a small number of differentially expressed ECM proteins. Several factors including the quality of *postmortem* human samples and methods applied for sample dissociation and extraction may affect the protein extraction (145). Postmortem interval (PMI) is one of the important parameters in postmortem studies, particularly when evaluating postmortem tissue sample quality. There has been evidence that a prolonged PMI causes protein breakdown, which substantially reduces the amount of detectable protein during subsequent tissue processing (145). However, if the autopsy is taken as soon as possible (PMIs <22 h), it was demonstrated that the protein integrity will be retained, which is consistent with the majority of the articles included in our review work. In most of the articles the age, the gender and the PMI were reported to be matched/controlled. Therefore, other variability between individual patients and overall health at the time of death may be more likely here (145, 146).

In conclusion, limited relevant studies were accessed from our search indicating that much work still remains to be done to better understand the roles of the ECM in neurodegeneration and PD. Our work summarized proteomic and transcriptomic studies of ECM genes and proteins that are dysregulated in PD. From the collective evidences, we observed that, although the current knowledge on the involvement of aberrant ECM proteins in PD is still in its infancy, it is clear that changes in expression of the ECM macromolecules play important roles in PD. Annexins, collagen VI, versican, and tenascins were the widely reported differentially expressed proteins whereas ECM-receptor interaction, focal adhesion, cell adhesion molecules and cell adhesion together with the integrin signaling pathway and individual integrin genes were commonly dysregulated at the transcription level.. These ECM components and pathways are potential sites to be investigated, validated and used as a drug targets for PD treatment.

## Data Availability

All data produced in the present study are available upon reasonable request to the authors

## Acknowledgments

The authors would like to thank the Israel Science Foundation (ISF grant 1994/21 and 3252/21) and Zuckerman (Zuckerman STEM leadership program) for funding and support to Dr. Shani Stern

